# High Resolution analysis of Transmission Dynamics of Sars-Cov-2 in Two Major Hospital Outbreaks in South Africa Leveraging Intrahost Diversity

**DOI:** 10.1101/2020.11.15.20231993

**Authors:** San Emmanuel James, Sinaye Ngcapu, Aquillah M Kanzi, Houriiyah Tegally, Vagner Fonseca, Jennifer Giandhari, Eduan Wilkinson, Benjamin Chimukangara, Sureshnee Pillay, Lavanya Singh, Maryam Fish, Inbal Gazy, Khulekani Khanyile, Richard Lessells, Tulio de Oliveira

**Author notes:** **Correspondence:** Tulio de Oliveira ( and). **Authors contributed equally:** San Emmanuel James and Sinaye Ngcapu.

## Abstract

Severe acute respiratory syndrome coronavirus 2 (SARS-CoV-2) causes acute, highly transmissible respiratory infection in both humans and wide range of animal species. Its rapid spread globally and devasting effects have resulted into a major public health emergency prompting the need for methodological interventions to understand and control its spread. In particular, The ability to effectively retrace its transmission pathways in outbreaks remains a major challenge. This is further exacerbated by our limited understanding of its underlying evolutionary mechanism. Using NGS whole-genome data, we determined whether inter- and intra-host diversity coupled with bottleneck analysis can retrace the pathway of viral transmission in two epidemiologically well characterised nosocomial outbreaks in healthcare settings supported by phylogenetic analysis. Additionally, we assessed the mutational landscape, selection pressure and diversity of the identified variants. Our findings showed evidence of intrahost variant transmission and evolution of SARS-CoV-2 after infection These observations were consistent with the results from the bottleneck analysis suggesting that certain intrahost variants in this study could have been transmitted to recipients. In both outbreaks, we observed iSNVs and SNVs shared by putative source-recipients pairs. Majority of the observed iSNVs were positioned in the S and ORF1ab region. AG, CT and TC nucleotide changes were enriched across SARS-COV-2 genome. Moreover, SARS-COV-2 genome had limited diversity in some loci while being highly conserved in others. Overall, Our findings show that the synergistic effect of combining withinhost diversity and bottleneck estimations greatly enhances resolution of transmission events in Sars-Cov-2 outbreaks. They also provide insight into the genome diversity suggesting purifying selection may be involved in the transmission. Together these results will help in developing strategies to elucidate transmission events and curtail the spread of Sars-Cov-2

## Introduction

The emergence of a novel coronavirus, severe acute respiratory syndrome coronavirus 2 (SARS-CoV-2) in the Wuhan district of China, and its rapid spread and increasing death toll resulted in a public health emergency of international concern in just under two months (Zhu et al., 2020, WHO, 2020). In South Africa, the first officially diagnosed case of SARS-CoV-2 was reported on March 5, 2020. The strict public health mitigation strategies and non-pharmaceutical interventions played a critical role in controlling the COVID-19 pandemic in South Africa, lowering the new cases reported each day to approximately 1500 (as of October 23, 2020; https://www.nicd.ac.za/diseases-a-z-index/covid-19/surveillance-reports/). While South Africa marks a significant decline in SARS-CoV-2 cases, understanding the patterns of transmission and underlying selection pressures within and between susceptible host populations remain critical to control and prevent further outbreaks.

Phylogenetic inference has mainly been used together with epidemiological investigations to elucidate transmission events and retrace transmission pathways (He et al., 2020, Lauring, 2020, Bajaj and Purohit, 2020, Guo et al., 2020), however, reports of these investigations using phylogeny only represent the dominant viral lineage and thus provide limited resolution in transmission analyses (Mavian et al., 2020). Whole genome analyses integrating withinhost diversity have been proposed as a better alternative in order to capture the complete genetic architecture including divergent, transient and deleterious variants in viral populations represented within a given host(Sanjuan et al., 2004). Indeed, several studies have already revealed the existence of large numbers of withinhost viral populations in SARS-CoV-2 (Wolfel et al., 2020, Shen et al., 2020, Lythgoe et al., 2020, Butler et al., 2020). Genomic and quasispecies analyses showed large numbers intra-host nonsynonymous mutations in nasopharyngeal and oropharyngeal swabs of SARS-CoV-2 positive cases (Zhou et al., 2020a, Siqueira et al., 2020). Intrahost single nucleotide variants (iSNVs) were were also observed in bronchoal-veolar lavage fluid samples of of 8 SARS-CoV-2 patients(Shen et al., 2020). And Wang et al. (2020) reported samples carrying the same consensus sequences exhibiting different iSNVs. Interestingly, Shen et al. (2020) could not confirm the transmission of iSNVs across two confirmed source-recipient pairs in the Wuhan area, suggesting either the effect of strong purifying selection or stochastic occurrence of variants within the host(Shen et al., 2020). It therefore remains unclear whether intrahost single-nucleotide variants can enhance resolution of transmission dynamics of SARS-CoV-2.

Furthermore, it has been reported that majority of variants identified tend to be a result of stochastic effects and selection pressure thus without any functional effect. Others, however, in combination with the host genetic profile, can produce viral genotypes with altered pathogenicity and improved host adaptation (Lucas et al., 2001, Gojobori et al., 1990). It is therefore important to understand the genomic diversity of SARS-CoV-2, the implication of identified mutations and evolutionary pressure on the virus beyond interhost single nucleotide variantion level. This is particularly due to the fact that little is currently known about the evolutionary dynamics of the SARS-CoV-2 owing to the differential evolution of the virus across distinct populations.

Leveraging our existing genomic and epidemiology data (Giandhari et al., 2020, Pillay et al., 2020) from two well characterized nosocomial Sars-Cov-2 outbreaks in healthcare settings with advances in whole genome analysis, we performed indepth analysis of the transmission patterns and pathways of Sars-Cov-2. We further describe in detail the distributions of inter and intrahost single-nucleotides variants and characterize evolution of the virus across the SARS-CoV-2 genome. We show that combining withinhost diversity and bottleneck estimation presents a major tool to retrace the pathway of viral transmission in an Sars-Cov-2 outbreaks.

## Methods

### Sample Collection and Detection of SARS-CoV-2

Data analyses in this study comprised of 109 SARS-CoV-2 cases from two nosocomial outbreaks from within healthcare settings in the Kwazulu-Natal province of South Africa. The first outbreak was a 4 weeklong outbreak where phylogenetic analysis supported the hypothesis of a single introduction leading to multiple chains of transmission within the hospital. The second outbreak occured a little later but still at a time of relatively limited community transmission within in a specific section of the hospital. The clustering was supported by phylogenetic analysis, but the precise introduction of the virus was not clear, and the precise chains of transmission were not elucidated. The CH1 outbreak has been further described in detail by (Giandhari et al., 2020, Pillay et al., 2020) and in a genomic epidemiology report by Lessels et al. (2020) (https://www.krisp.org.za/news.php?id=421). Clinical specimens from the one hundred and nine symptomatic cases were tested COVID-19 and infection status was confirmed using comparative real-time qPCR

### Amplification and Illumina MiSeq Sequencing

A 400 base pair (bp) product with 70bp overlap that covers the SARS-CoV-2 genome was amplified by multiplex PCR, using primers designed on Primal Scheme (http://primal.zibraproject.org/) as previously described (Giandhari et al., 2020). PCR products were purified using AmpureXP purification beads (Beckman Coulter, High Wycombe, UK) and quantified on a Qubit Fluorometer using Qubit dsDNA High Sensitivity assay according to the manufacturer’s instructions. Sequencing reads were generated using libraries prepared by Illumina® TruSeq® Nano DNA Library Prep kits and spiked with 1% PhiX and loaded onto a 500-cycle MiSeq Nano Reagent Kit v2 nano v2 Miseq reagent kit and run on the Illumina MiSeq instrument (Illumina, San Diego, CA, USA) as previously described (Pillay et al., 2020)

### Assembled sequence and iSNV analysis

Consensus genomes used in the study were generated following a standard bioinformatics protocol available at (protocols.io/private/CD7108C5B73211EA89B10A58A9FEAC2A) (Cleemput et al., 2020). We used a modified version (NC_045512.3) of the NC_045512.2 reference strain from the Swiss Institute for Bioinformatics (SIB) available at https://github.com/jsan4christ/Sars-Cov2-Refs/blob/master/NC_045512.2-sib.gb, to call and annotate variants. We used LoFreq v.2.1.5 (Wilm et al., 2012) to call variants from the mpileup files generated using samtools software (version 1.8) (Li et al., 2009). Only variants called at a sequencing depth of ≥10 reads and with a minor allele frequency of ≥5% were considered for further downstream analysis. Positions with more than one minor allele were filtered out to minimize the false discovery of variants. Variants not found in coding regions of the genome were also filtered out. Variants were then annotated using snpEff v. 4.5 (Cingolani et al., 2012b) and variation effects filtered using SnpSift v.4.3t (Cingolani et al., 2012a).

#### Genetic Diversity

We estimated genetic diversity and the non-synonymous to synonymous diversity ratios using SNPGenie (Nelson et al., 2015). The between-group mode of SNPGenie was used to evaluate the mean *d*N and *d*S between the outbreaks while default mode (SNPGenie.pl) was used to estimate diversity for sites in protein-coding sequences of each sample from the associated VCF reports.

#### Transmission Analyses

To determine the iSNVs shared between related cases and the possibility of transmission, we compared variant calling results of putative source-recipient pairs of samples. For CH1, we leveraged known source-recipient pairs while all samples were treated as a potential source and potential recipient in the CH3 outbreak due to a lack of putative source-recipient links. We then applied the betal-binomial model, as implemented in the BB Bottleneck software (Sobel Leonard et al., 2017), to estimate the founding population of the virus in the recipient. The beta binomial model is superior to the mutation counting method in its ability to account for variant calling thresholds and stochastic viral replication dynamics within the recipient hostv(Sobel Leonard et al., 2017). To generate the source-recipient frequencies required as input to BB bottleneck, we followed the approach described by (Sapoval et al., 2020). Briefly, for the source, where the alternative allele frequency was > 0.5, the reference allele was taken as the minor allele and the frequency as 1 – AF (Allele Frequency). We considered the minor allele from the source as the reference or alternate allele, for the recipient,. In case the minor allele does not exists, the position was assigned a frequency of 0. Finally, all positions had to be supported by atleast 10 reads to be considered both for the source and recipient.

## Results

This study included a total of 109 SARS-CoV-2 cases. Clinical characteristics of individuals are reported in Supplementary Table 1. The 109 cases were further categorized by outbreak as CH1 (35/109, 32%) and CH3 (74/109, 68%). Of the 35 samples collected from CH1 that were available for our analysis, 13 (37.1%) had putative transmission linkages that were phylogenetically supported(Figure 1a). Samples from the beginning of the CH3 outbreak (24/74, 32.4%) that were part of the epidemiologically investigation were grouped by department and social networks (Figure 1b). Phylogenetic analysis further grouped these samples into a single large cluster with four connected outliers and three samples unrelated to the major cluster (Figure 1c).

**Figure 1.**
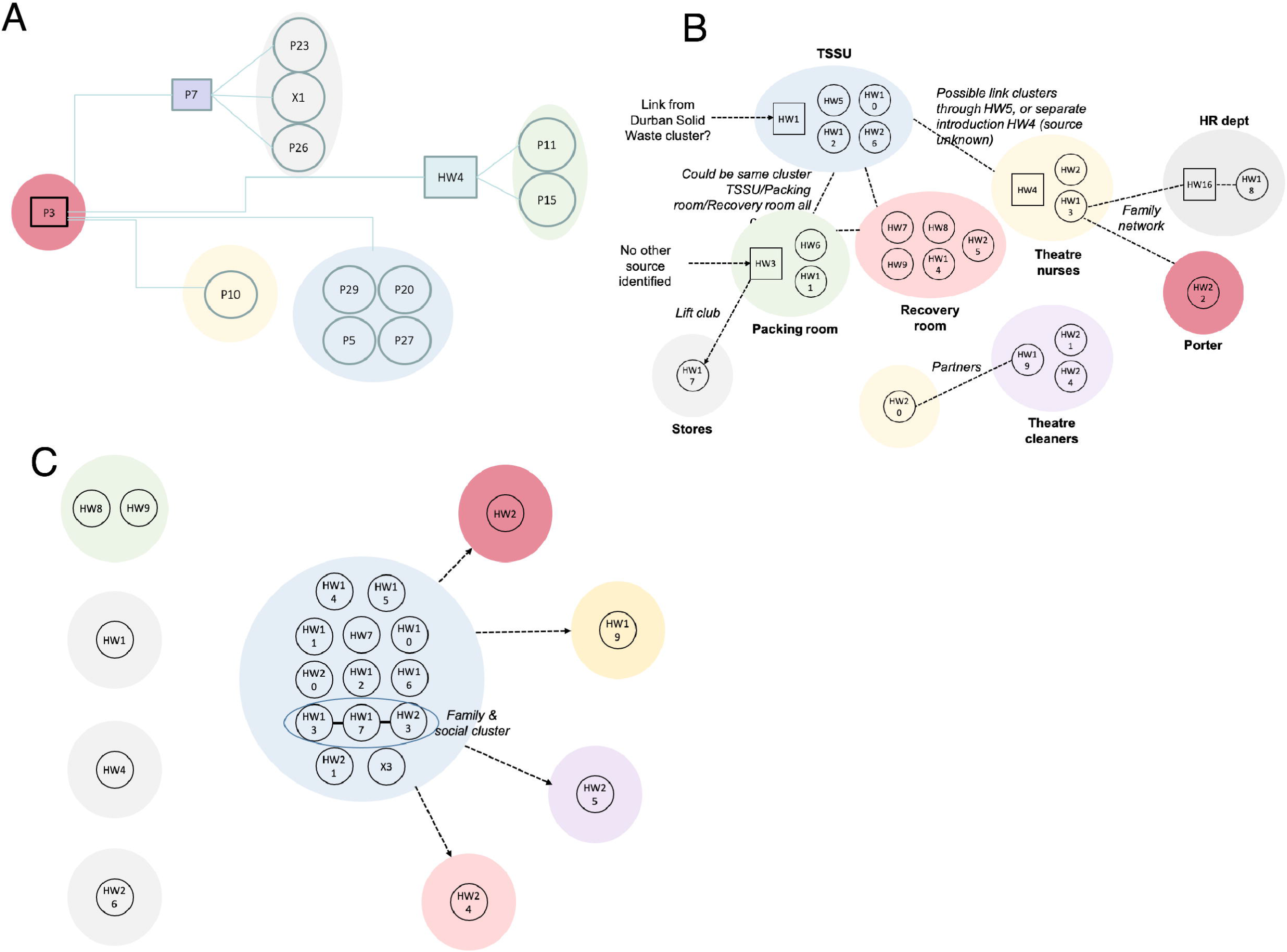
Transmission maps showing epidemiological and phylogenetic linkages of SARS-CoV-2 infections in different hospital outbreaks in Durban. (A) Schematic of networks of known sources of transmission (squares) and patient that became infected (circles) in the CH1 outbreaks. (B) Epidemiological distribution of CH3 patients by work department and social clusters within the hospital to narrow infective space. Patient HW7 is suspect to be the source of the first introduction. (C) Phylogenetic distribution of CH3 patients into a single large cluster. Attached samples closely related to the mainly cluster while non-attached samples had no relationship with the other samples. Squares indicate potential source of transmission and circles represent patient that became infected in the outbreaks.

### Allele frequencies and mutational landscape in SARS-CoV-2 genes

All 109 whole-genome sequences analyzed yielded near full-length genomes with coverage greater than 90% and the average read depth ranged from 158.71 – 5046.56 (Supplementary Table 2). The read depth did not have any impact on the minor allele frequency. In total, 2552 iSNVs were identified across 109 CH1 and CH3 samples at MAF >5% (Figure 2A). The majority of these nucleotide changes occurred in CH1 than CH3 samples, with the exception of GC (42 vs 49) and GT (111 vs 130). We also observed 940 SNVs, with different mutational patterns to iSNVs. These include CT (n= 412), AG (n=201), GA (n=100) as the most prevalent. The frequency of the SNVs was high in CH3 samples compared to CH1. In terms of location in the SARS-CoV-2 genome, a large fraction of iSNVs and SNVs were found within the S gene and non-structural proteins of the ORF1ab gene, specifically in the NSP3 protein (Supplementary Table 3).

***Table 1.*** *Summary of iSNVs present at >5% frequency in 109 SARS-CoV-2 genomes*

**Figure 2.**
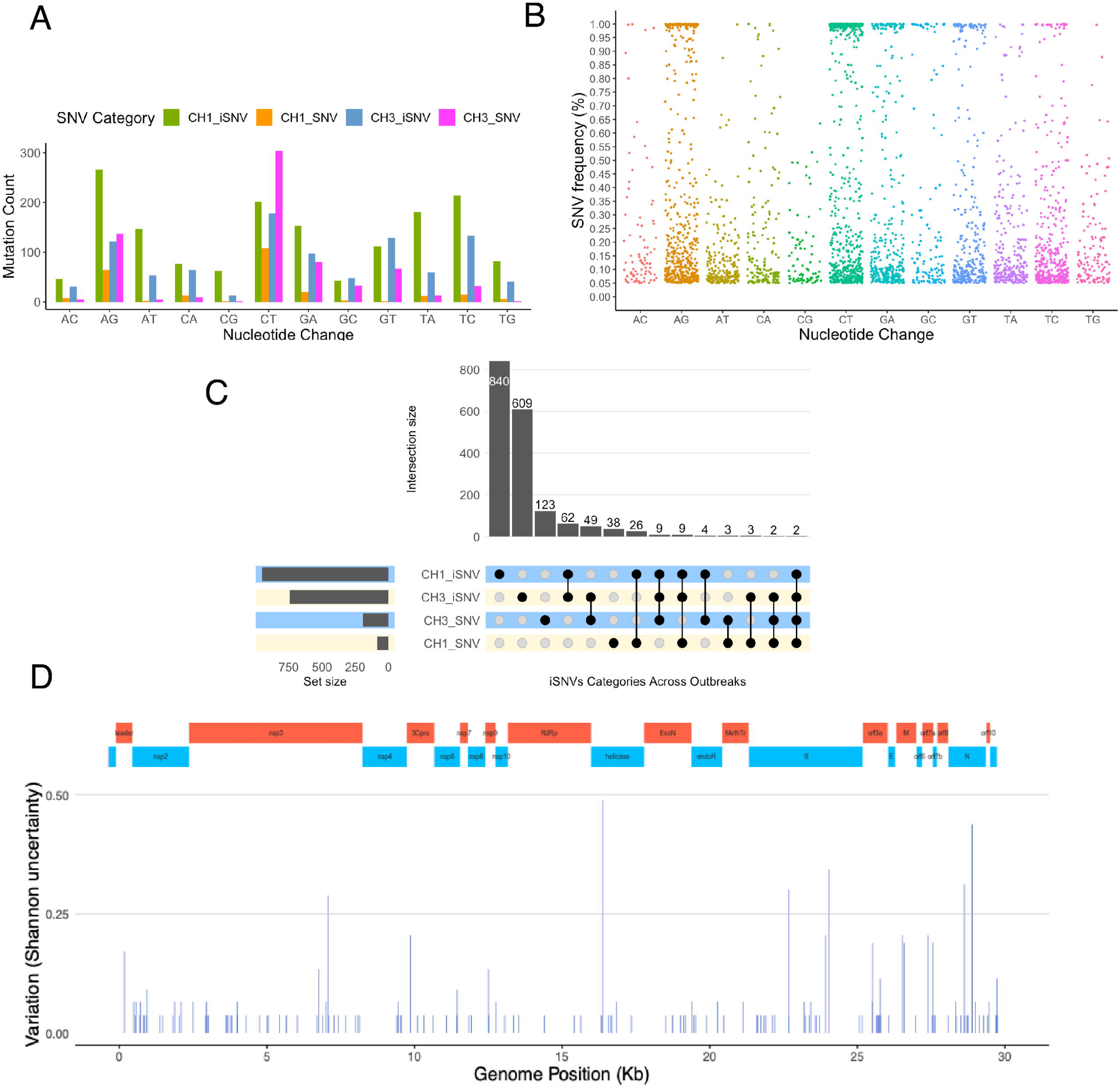
Overview of general diversity of SARS-CoV-2 genomes from South African patients. (A) Nucleotide changes in SARS-CoV-2 genomes. (B) Distribution of variant frequencies across nucleotide changes. (C) The upset plot shows the distribution of iSNVs and SNVs across the outbreaks. The vertical bar chart shows the size of the intersection and the black dots and lines show the combination of iSNVs and SNVs. The horizontal bars shows the unconditional frequency count of variants across within each group. (D) Sequence variability detected in SARS-CoV-2 overlaid with major structural protein coding regions in the genome.

The 3492 iSNVs and SNVs were further categorized as 2376 non-synonymous, followed by 997 synonymous, and 119 nonsense (stop lost/gained) (Table 1, Supplementary Table 3). Of the observed iSNVs, the majority of non-synonymous (1121 vs 652), synonymous (392 vs 278) and nonsense (71 vs 63) were found in the CH1 samples compared to CH3 samples. In contrast, a large fraction of non-synonymous (421 vs 182), synonymous (264 vs 63) SNVs were found in the CH3 than in CH1 samples, with the exception of CH1 nonsense SNV (7 vs 3). The iSNVs were distributed in 11 protein-coding viral genes with variable frequencies, with ORF1ab gene harbouring the majority of non-synonymous and synonymous variants compared to other genes. Figure 2B shows that the most frequent iSNVs were found in the non-structural proteins (NSP)8 (A12240G in 24/109 samples), NSP14 (G18181T in 22/109 samples), S (T21667G in 21/109 samples), NSP9 an NSP13 had A11556T and A13003G in 20 samples, respectively. We also observed a high density of polymorphic SNVs A23403G (109/109 in S gene), C14408T (109/109 in NSP12), C3037T (104/109 in NSP3) in the analyzed samples. Other genes (E, M and N) and proteins (NSP10, NSP11, NSP16, NSP5, NSP4, NSP7, ORF10, ORF6, ORF7a, ORF7b and ORF8) were well conserved, with frequencies <5% for both iSNVs and SNVs.

### Transmission of consensus mutations between source-recipient pairs in the first hospital outbreak

Here, we used consensus mutations to explore the transmission dynamics of SARS-CoV-2 within and across samples of thirteen in-hospital patients and healthcare workers (HW). In our report into a nosocomial outbreak of SARS-CoV-2 infections at one of the private hospitals in Durban, phylogenetic inferences showed that inpatient-3 (P3, source) infected by the index patient sustained the chains of transmission generating secondary clusters of recipients including HW4 and P7. These findings were supported by common mutations (C241T, C3037T, C14408T, A23403G) found in the viral consensus sequences of (P3, Cluster1) and its putative recipients (P5, P7, P20, P27 and P29) (Table 2). In addition, P5, P20, P27 and P29 also developed an additional mutation C16376T. Similar mutations were found in the secondary clusters 2 (HW4), and 3 (P7) with additional mutations found in consensus sequences of patient P26 (A16561C) and X1 (C2997T) in cluster 3. Development of additional mutation could be attributed to multiple transmitted strains, selection pressure within the host or some positions being prone to mutations.

***Table 2.*** *Common consensus mutations shared between putative source-recipient pairs in the CH1 outbreak*

### Transmission dynamics of shared intrahost variants between samples

Here, we investigated whether minor alleles with a frequency of 5% or higher observed varied between the epidemiologically infered source and recipients and whether these can be indicative of transmission events within the CH1 and CH3 hospital outbreaks. From the thirteen CH1 samples analyzed, we observed 615 iSNVs. The CH1 iSNVs consisted of 442 non-synonymous, 146 synonymous and 27 nonsense variants (Supplementary Table 3). We assessed the evidence for the transmission of shared iSNVs between samples. We found that HW4 (source) was most likely to have transmitted eleven iSNVs to the recipient P11 and three of which were positively selected for and later established as SNV (Figure 3A, Table 3). In addition, P7 potentially passed on nine iSNVs to P23 (Figure 3B) and sixteen to X1. Of those sixteen, three were later established as SNVs (Figure 3C). Furthermore, P3 shared seven iSNVs with P5 (Figure 3D) and P7 (Figure 3E). P3 was likely to have transmitted four iSNVs to P10, one of which later established as an SNV (Figure 3F). There were eleven shared iSNVs between P3 and P20, with two later established as SNVs (Figure 3G). P3 shared nine iSNVs with P29, with three later established as SNVs (Figure 3H). In contrast, there was no evidence of shared iSNVs between P3 (source) and recipient P27. Similarly, sources HW4 did not share any iSNVs with the recipient P15. Thirty five (i)SNVs were shared by two or more samples (Supplementary Figure S1). These findings are consistent with the results from the bottleneck analysis, albeit a much smaller founding population estimated to range from one to three (Table 3). Based on these findings, we see that three of the eleven pairs are were not directly related and could potentially have alternate infection sources.

***Table 3.*** *Shared iSNVs and bottleneck estimates in CH1 outbreak putative source-recipient pairs*

**Figure 3.**
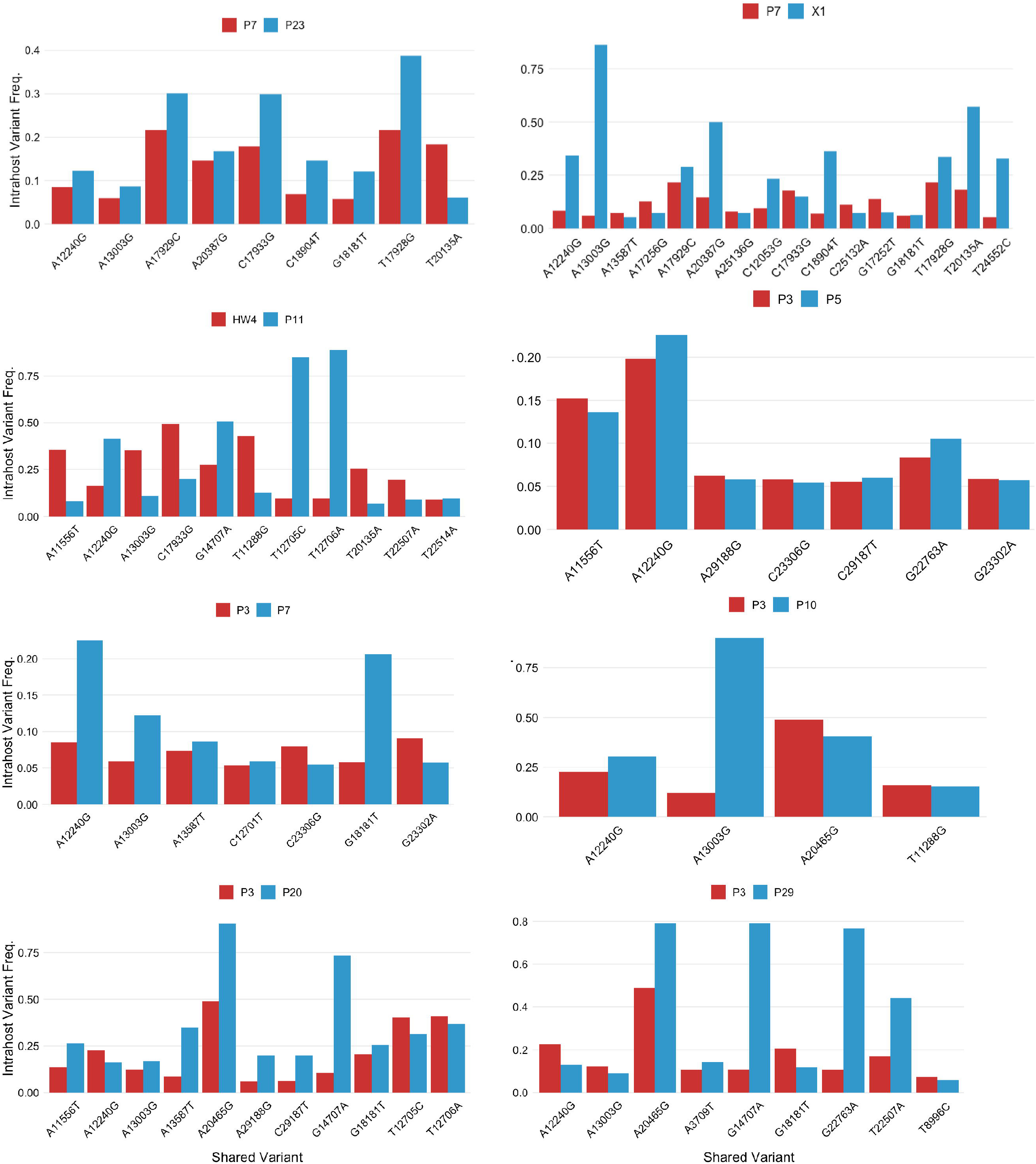
Transmission dynamics of shared intrahost variants between samples. Each plot shows shared iSNVs between putative donor (red)/recipient (blue) pairs as evidence for transmission. (A) Presence of shared iSNVs between P7, P23 and (B) X1. (C) shows shared iSNVs, which later established as SNVs in the recipient, between HW4 and P11. (D) P3 shared iSNVs with P5, and (E) P7. P3 iSNVs, which later established as SNVs, were also shared with (F) P10, (G) P20 and (H) P29.

We also explored CH3 samples for co-occurring iSNV as indicative of transmission events in the population. Since there were no putative source-recipient links in CH3 samples, we leveraged pairwise sample comparison and bottleneck estimations to explore putative transmission events within and between departments and social networks. Thus we generated 5402 potential source-recipient pairs from seventy four patients and explored the presence of source iSNVs as iSNVs or SNVs in the respective recipient. Among observed iSNVs, the majority of pairs (4151/5402) had no shared iSNVs while 1251/5402 pairs had one or more iSNV (Figure 4A). In terms of nucleotide position of shared iSNVs, we observed two positions 21665 and 21667 found in 227 and 440 pairs that exhibited strong signals for shared iSNVs, respectively (Figure 4B). Other noticeable positions with strong signals for shared iSNVs were 25653 and 25666, respectively. In CH3 samples, HW7 was identified as the most likely source of infection by the epidemiological investigation supported by phylogenetic inference. However, this was not supported by number of shared iSNVs and bottleneck estimates. HW7 shared not more than 2 iSNVs with the cases that were clustered together by phylogeny. (Supplementary Table 4)(Figure 1C). HW19 and HW20 were from the same household and shared upto eighteen iSNVs. They also had bottleneck estimates of 3 in the direction HW19 to HW20 and estimated size of 6 when swapped around (HW20 to HW19), suggesting that HW20 was the more likely source of infection to HW19. HW25 also shared 17 and 20 iSNVs with HW19 and HW20 suggesting possible transmission of the virus from HW19 to HW25. Overall, these findings suggest that certain intrahost variants in this study could have been transmitted to recipients; however, most intrahost variants occurred after transmission.

**Figure 4.**
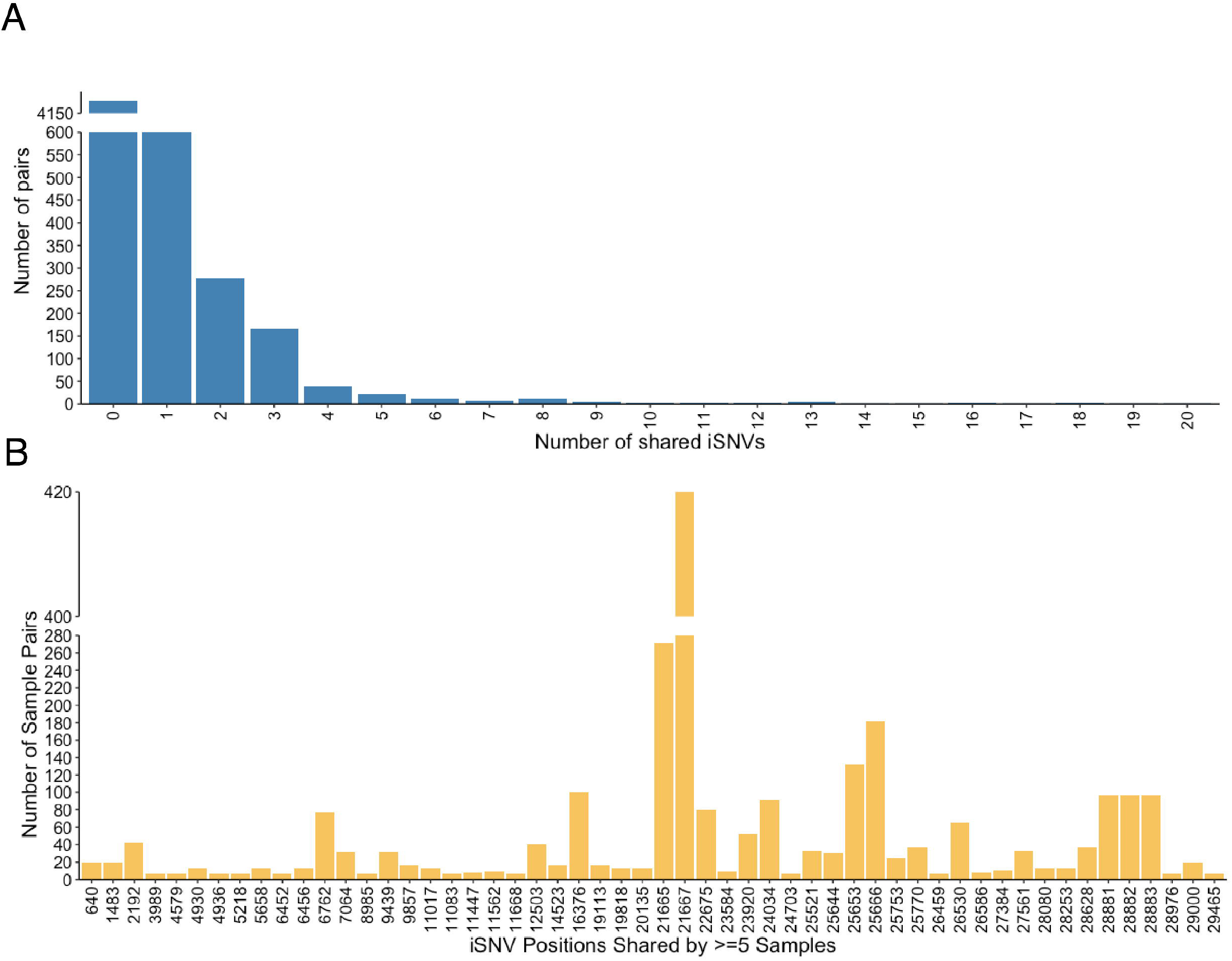
Putative iSNVs transmission events amongst CH3 samples. (A) Bars show distribution of the number of shared iSNVs amongst CH3 pairs and (B) at given nucleotide positions. Majority of pairs had no shared iSNVs while positions 21665 and 21667 exhibited strong signals for shared iSNVs.

### Variant Prioritisation and Pervasion Across Outbreaks

Most of the variants expressed in viral sequences usually do not confir any advantage ot the virus therefore get purged. A small set of likely advantageous mutations are however positively selected for and therefore established as clade or lineage defining. Using the upset plot (Figure 2C), we further captured the intersections between iSNVs and SNVs in the CH1 and CH3 datasets to identify frequently and universally occurring iSNVs. 10 mutations occurred in CH1 and CH3 independently. We also found 37 and 63 iSNVs that occurred as SNVs in atleast one CH1 and CH3 sample, respectively. Variants T20135A and A14523G occurred as iSNVs and SNVs in both outbreaks. Figure 2D shows the Shannon diversity observed along the SARS-CoV-2 genome. iSNVs progressing to SNV are likely of interest for further research.

### Selection pressure in the SARS-CoV-2 genome

To assess impact of selection pressure on transmission dynamics and establishment of the virus within and between hosts, first, we estimated the nucleotide diversity (π) and genetic complexity (S_*n*_) of CH1 and CH3 samples using pairwise comparison for each protein-coding gene. We observed median diversity (median = 0.00004) within the samples analyzed with diversity varying between 0 and 0.001. We then analysed diversity by comparing the non-synonymous to synonymous ratios (πN/πS) to determine the nature of selection pressure affecting the sample’s nucleotide diversity from both outbreaks. CH3 samples exhibited a low mean Sn (KS test: p-value < 10^−8^) and mean π (KS test: p-value < 10^−8^) compared to CH1 samples (Figure 5A & B). We further compared the non-synonymous and synonymous diversity ratios between the two outbreaks. CH1 ratios were significantly higher (median πN/πS: 0.0004) than CH3 ratios (median πN/πS: 0.00003 and KS test: p-value < 6.8^−4^) (Figure 5C & D). We classified our samples according to selection pressure on the viral population withinhost. In CH1, 8% (3/35) of the samples exhibited neutral selection, 31% (11/35) were under positive selection while the majority, 60% (21/35), were under purifying selection. We observed a similar trend in CH3 with 8% (6/74) under neutral selection, 21% (16/74) under positive selection and 70% (52/74) under purifying selection. Diversity analysis of ORFs within the genome revealed positive selection in N, ORF1ab, ORF3a and S (Figure 5E). Overall, our diversity results correlate with the high conservation (>90%) observed between SARS-COV-2 genomes (Khan et al., 2020). We also applied the between-group mode of SNPGenie to further compare the diversities between CH1 and CH3 at a consensus level. The highest dN/dS ratios between group were observed in N (9.12) followed by S, ORF7a (>3), ORF3a (>2) and ORF9b (>1) (Table 4). Overall, there was no significant difference between the nature of selection pressure observed across the two outbreaks (KS test, p-value = 1). However, the genome-wide πN/πS ratios suggest that SARS-COV-2 intrahost and interhost diversities in the population are mostly under purifying selection.

***Table 4.*** *dN/dS results from between group diversity analysis of CH1 and CH3 outbreaks*

**Figure 5.**
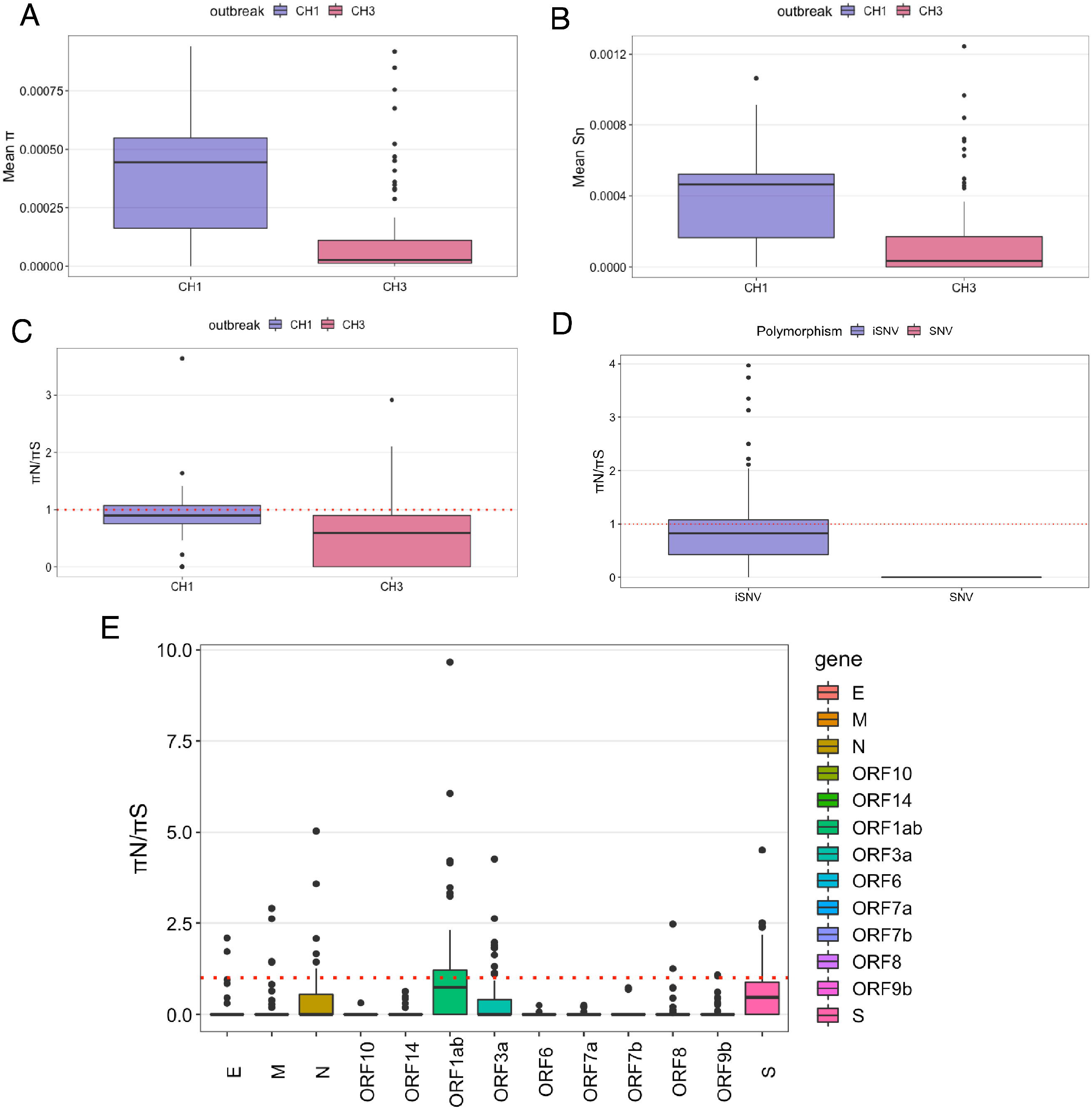
Estimated nucleotide diversity and genetic complexity in SARS-CoV-2 genomes of of CH1 and CH3 populations. (A) Box plots show mean nucleotide (π) diversity, and (B) mean (S_n_) genetic complexity (selection pressure) of CH1 and CH3 samples. (C) Ratio of non-synonymous to synonymous diversities across CH1 and CH3 samples and (D) ratio of non-synonymous to synonymous diversities for iSNVs and SNVs across the outbreak. (E) Genome-wide diversities (πN/πS ratios) observed in each gene product or ORF

## Discussion

In the present study, we assessed the utility of intrahost diversity in the elucidation of transmission events between hosts. We confirmed that our analysis could bring better resolution to transmission events. The synergy of intrahost diversity and bottleneck estimation brough more support to the introduction of virus and direction of transmission between source-recipient pairs in both outbreaks. Of the 11 source-recipient pairs from CH1, eight shared intrahost variants, suggesting person to person transmission of intrahost variants was possible in SARS-COV-2. In addition, these transmission patterns were further supported by bottleneck analysis, with estimated bottleneck sizes between 1 and 3. Likewise, three pairs without any shared iSNVs also lacked bottleneck supports suggesting that infact, these pairs might not be related as initially implied by the epidemiological investigation and phylogenetic inference. Futhermore, using shared iSNVs and bottleneck estimates between CH3 source-recipient pairs, we brought additional support to the epidemiologically infered transmission patterns that were originally not clear from phylogenetic analysis. We show that indeed HW7 is a probable source of introduction of the virus, but also highlight HW10, HW11, HW15 and HW16 as possible candidates. We also show that HW20 most likely infected HW19 using bottleneck estimates. The narrow transmission bottleneck observed in these outbreaks may be attributed to a small number of virions that crossed the host cell barrier and established infection or deleterious stochastic dynamics within the respiratory tract (Wang et al., 2020). Another reason for narrow transmissions could be attributed to the merging or adaptive evolution of the virus, which could potentially affect the virus’s antigenicity and pathogenicity (Berngruber et al., 2013). Some samples however, had very high bottleneck estimates above 50 virions (mean deviation range of 57 to 1000).

In order to understand the impact of selection pressures on the patterns of variation represented in the transmission events, we assessed the frequency and diversity of inter- and intra-host variants identified. We found an excess of AG, CT, TC and GA mutations in both iSNVs and SNVs across all samples analyzed. In addition, we found that ORF1ab gene harboured the majority of non-synonymous and synonymous variants compared to other genes. Between the outbreaks, CH3 samples has a low mean Sn and mean π compared to CH1 samples. Furthermore, there was evidence of unique or shared iSNVs between the samples in both outbreaks. In both outbreaks, the mutational spectrum of new mutations seems enriched in AG, CT, TC and GA mutations. S gene had the largest fraction of iSNV and SNV mutational patterns, followed by ORF1a protein such as NSP3 and NSP12. Particularly, S gene was dominated by GA and AT mutations, NSP3 by CT, GA, TA and TC mutations and NSP12 had CA dominance, both for iSNVs and SNVs. Mutations GT in the NSP14; GC and TG in the S were also common in the SARS-COV-2 genome. These findings are consistent with previous studies that found enrichment of CT mutations in the ORF1a gene (Di Giorgio et al., 2020, Sapoval et al., 2020). It has been suggested that the CT mutation enrichment in the SARS-COV-2 genome is likely driven by host response to counter the virus through the APOBEC and ADAR deaminase activity. These studies also note that mutation changes in AG and TC in the SARS-COV-2 genome were mediated by the actions of ADARs while GA mutations were derived from APOBEC-mediated C-to-U deamination (Di Giorgio et al., 2020, Sapoval et al., 2020, Roth et al., 2019, Porath et al., 2014). These findings suggest that S gene and ORF1ab gene had higher substitution rates and observed changes of the SARS-COV-2 genome might be hyper-edited by APOBEC and ADAR.

When assessing the genomic diversity of SARS-CoV-2 within the two outbreaks, we found that ORF1ab, N, ORF3a and S were relatively diverse (Figure 5E). Diversity analysis between the two the two oubreaks further showed signals of strong positive selection in the N gene, S and ORF7a. Positive selection was also seen in ORF3a and ORF9b (Table 4). On the other hand, ORF1ab, OFR14 and M exhibited purifying selection suggesting that these gene products did not undergo adequate immune pressure resulting in amino acid changes. This is consistent with other studies that reported strong evolutionary pressure in N, S and ORFs regions (Zhou et al., 2020b, Cagliani et al., 2020) causing them to evolve faster. It has been shown that ORF1ab encodes proteins involved in SARS-CoV pathogenesis, virulence, virus-cell interaction and alterations of virus-host responses by modifying cellular signaling and gene expression (Graham et al., 2008). ORF1ab has also been reported to encode 16 non-structural proteins that are highly conserved (Khan et al., 2020). ORF3a, localized in the Endoplasmic reticulum (ER) and Golgi region, is a transmembrane protein responsible for activating the PKR-like ER kinase signaling pathway to protect viral proteins against ER-associated degradation. Rapid accumulation of non-synonymous mutations in the ORF3a gene could increase the fitness of the viral population and help to escape host immune response, making it hard to eliminate and promoting viral spread (Issa et al., 2020, Minakshi et al., 2009).

In summary, we showed that combining withinhost diversity and bottleneck estimates can bring greater resolution to transmission analyses by providing insights into both chains of infection and direction of transmission. We also showed a complex landscape of intrahost diversity and evolution of SARS-CoV-2 after infection, with a predominance of purifying selection explaining the small number of shared intrahost variants inspite of larger estimated founding population. This study therefore enhanced our understanding of potential viral transmissions within and across SARS-CoV-2 cases and shed light on the use of intrahost variants and bottleneck estimates to retrace the pathway of viral transmission in a population. Results obtained from this study strongly support additional research on the role of the observed intrahost variants in SARS-COV-2 antigenicity and pathogenicity to shed light on biological mechanisms driving the rapid spread of SARS-CoV-2 and complicated disease progression.

## Data Availability

All data is public available at GISAID and short reads at the Short Read Archive of NCBI.

https://www.gisaid.org

https://www.krisp.org.za/ngs-sa/ngs-sa_open_science_and_sharing_data_rapidly_but_believe_that_research_should_be_fair/

## Acknowledgments

We wish to thank all laboratory personnel that have worked to genotype SARS-CoV-2 samples. This study was funded by a research Flagship grant from the South African Medical Research Council (MRC-RFA-UFSP-01-2013/UKZN HIVEPI), by the Technology Innovation Agency and the Department of Science and Innovation and by National Human Genome Research Institute of the National Institutes of Health under Award Number U24HG006941. H3ABioNet is an initiative of the Human Health and Heredity in Africa Consortium (H3Africa). The content is solely the responsibility of the authors and does not necessarily represent the official views of any of the funders.

## Data Availability

Viral consensus genomes reported in this article have been deposited at Global Initiative on Sharing All Influenza Data (GISAID) [All accessions in Supplementary Table 1] (Shu and McCauley, 2017) while the raw FastQ sequences have been deposited to the National Center for Biotechnology Information (NCBI) Sequence Read Archive (SRA) (Leinonen et al., 2011) (Project Accession No. PRJNA636748).

## Authors Contributions

SEJ, SN, RL and TdO conceived and designed the analysis and SEJ, SN, AMK, EW, RL and TdO performed the analyses. SEJ, SN, AMK, HT, VF, JG, BC, SP, LS, MF, IG, EW, KK, RL and TdO have contributed to the interpretation and discussion of the results and writing of the manuscript.

